# Facilitators and barriers for healthcare workers’ adherence to the national nutritional guidelines for people living with HIV in Dar-Es-Salaam: A mixed-method study

**DOI:** 10.1101/2024.08.13.24311936

**Authors:** Felistar Mwakasungura, Rebecca Mkumbwa, Bruno Sunguya

## Abstract

The dual burden of HIV and malnutrition is a significant public health issue in Sub-Saharan Africa. In Tanzania, with an HIV prevalence of 4.9% among adults, malnutrition among People Living with HIV (PLHIV) remains high. This study evaluated adherence to nutritional guidelines for PLHIV in Care and Treatment Centers (CTCs) in Dar es Salaam and identified influencing factors. A mixed-method design was used among PLHIV, health facility administrators, and healthcare providers. Data were collected through observational checklists, structured questionnaires, and in-depth interviews. Statistical and qualitative analyses were performed to assess adherence and determinants. Among 478 participants who received care, only 19.46% were managed with fully adherence to the nutritional guidelines. Universally, anthropometric assessments were observed, however, micronutrient supplementation was minimal (1%). Higher education (AOR=5.08, p=0.019) and attendance at referral hospitals (AOR=8.23, p=0.032) positively influenced adherence. Conversely, nurse attendance (AOR=0.34, p=0.038), adequate staffing (AOR=0.31, p=0.010), and urban residence (AOR=0.47, p=0.009) negatively influenced adherence. Key facilitators included consistent training and supportive leadership, while barriers involved financial constraints and high staff turnover. The study reveals a significant gap in adherence to nutritional guidelines among PLHIV in Dar es Salaam, highlight the need for improved resource distribution, staff training, and patient education. Findings from this study aim to bridge the knowledge gap in adherence to nutritional guidelines for PLHIV, support evidence-based decision-making, and ultimately improve health outcomes.

## Introduction

Human Immunodeficiency Virus (HIV) remains a critical global health issue, with approximately 39 million people living with the virus as of 2022 [1]. Characterized by immune system suppression, HIV exacerbates nutritional challenges [2], particularly in low- and middle-income countries (LMICs). In Sub-Saharan Africa, which accounts for 66% of the global HIV population [3], the prevalence of malnutrition among People Living with HIV (PLHIV) is alarmingly high, at 23.7% [4]. This is compounded by a regional malnutrition rate of 21% [5].

The interaction between HIV and malnutrition creates a detrimental cycle [6]. HIV not only increases nutritional requirements [7], but also impairs the body’s ability to meet these needs due to symptoms such as oral thrush, painful swallowing, and nausea [8]. As a result, asymptomatic HIV-positive individuals need 10% more energy, while those with symptomatic HIV require 20-30% more energy compared to HIV-negative individuals [9]. This heightened energy demand, coupled with reduced food intake, further compromises the immune system, which is already weakened by HIV [7]. Nutritional care, when integrated with antiretroviral therapy (ART), can significantly improve health outcomes [10]. However, despite ART’s success in reducing HIV-related morbidity and mortality, nutritional concerns persist [11]. A severe shortage of healthcare workers skilled in nutritional care, particularly in Sub-Saharan Africa, exacerbates this issue [12]. As a result, about 10% of PLHIV in Care and Treatment Centers (CTCs) are undernourished, while others face issues of overweight and obesity [10].

In Tanzania, where HIV prevalence among adults stands at 4.9% [13], which 19.4% of PLHIV attending CTCs are malnourished [14]. The Tanzanian government has shown commitment to addressing both HIV and malnutrition through policies and guidelines such as the “National Guidelines for Nutrition Care and Support for People Living with HIV” and the “Health Sector HIV/AIDS Strategic Plan” [6]. These frameworks aim to improve nutritional care by integrating it into comprehensive health services, including assessment, education, counseling, and supplementation [6]. The “National HIV/AIDS Council Strategic Framework 2011-2015” emphasizes the importance of healthy eating and lifestyle as part of HIV care. The 2016 national guidelines for Nutrition Care and Support aim to coordinate nutrition programming, provide consistent information, and ensure that healthcare services include essential nutrition components such as assessment, education, counseling, and therapeutic feeding [6]. Despite these comprehensive strategies, the dual burden of HIV and malnutrition remains prevalent, impeding progress towards the 95-95-95 targets for HIV/AIDS elimination [15].

Evidence on adherence to these nutritional guidelines at the CTC level in Tanzania is scarce. The lack of data on guideline adherence and the factors influencing it continues to affect the quality of care and treatment outcomes for PLHIV, slowing progress towards effective HIV management and elimination goals. This study aims to evaluate the adherence to the National Guidelines for Nutrition Care and Support and to identify the facilitators and challenges associated with adherence in CTC clinics.

## Materials and methods

### Study design and area

The study was conducted across Care and Treatment Centers (CTCs) in Kinondoni, Temeke, Ilala, Ubungo, and Kigamboni districts of Dar es Salaam, encompassing urban and suburban areas. In 2023, more than 1.7 million people were living with HIV nationwide with approximately 4.7% of people living with HIV reside in Dar es Salaam [13], which encompassing nearly 10% of the country’s total population. This evaluation adopted the explanatory sequential mixed-method design, which had two phases which spanned between 29^th^ April and 27^th^ May 2024; A quantitative phase focused on level of adherence based on the observation of the researcher and patient feedbacks, whose results were used to build on the second phase of qualitative phenomenological design, targeting experiences of health care workers on providing care.

### Study population and sampling

*Quantitative phase;* A total of 485 PLHIV were approached for the study, of which 7(1.46%) refused to participate. Sample size was calculated using a cross-sectional study formula from Kish and Leslie (1965) with (p) of PLHIV’s adherence as 50%, and margin of error (E) set at 5%, and accounting for a 20% non-response rate. A multi-stage sampling method was employed to select 25 public health facilities with CTC clinics were selected using a multi-stage sampling method. From each of the facilities 19 participants were conveniently selected. Patients and treatment supporters who attended CTC clinics, health facility in-charges, doctors, nurses, and nutritionists who consented to participate were included. The exclusion criterion was clients experiencing acute medical distress at the time of data collection. *Qualitative Phase:* Thirteen healthcare providers were purposively approached and consented to participate in the study, ages ranged from 24 to 51 years, majority being male, five clinical officers, four nurses, three nutritionists, and two medical doctors, with their experience in working at CTCs ranging from one to eighteen years.

### Variables and measurements

Adherence level was the dependent variable, assessed using the Minimum Nutrition Package (MNP) per National Nutritional guideline, which contains a total of nine recommendations categorized into four core components: Nutrition Assessment, Education and Counseling, Therapeutic/Supplementary Feeding, and Referral/Follow-Up Services. Adherence was measured using a binary scoring system, with scores of 6 or more indicating adherence and scores below 5 indicating non-adherence, based on the Index Measuring Adherence to Complementary Feeding Guidelines [16, 17]. Independent variables including healthcare provider characteristics, health facility characteristics, resources and tools, training and supervision factors and patient characteristics, adopted from the Tanzania Demographic Health Survey (TDHS) [18], Tanzania Food and Nutrition Center (TFNC) [6], and Evaluation of facility level on NACS guide tool [19].

### Data collection and quality assurance

*Quantitative Phase*: A structured questionnaire was translated to Kiswahili. The Kiswahili version was uploaded on Kobo Collect. The questionnaire was tested on a pilot population at Tip top dispensary and Toagoma dispensary. The collected data were cleaned, processed, and securely stored in cloud storage to maintain confidentiality. This was complemented by Cronbach’s Alpha of 0.71. *Qualitative Phase:* In depth interview were conducted using tested semi structured interview guide, researcher recorded verbal and took notes on the non-verbal expressions.

### Data management and analysis

*Quantitative Phase*: Stata version 18 (Stata Corp, College Station, TX, USA) was used for analysis. Descriptive statistics were generated as mean and standard deviation for continuous variables, and frequencies and percentages for categorical variables. Bivariate and multivariate analyses were used to identify the significant determinants influencing adherence while adjusting for potential confounders. A threshold p-value of less than 0.05 was set for statistical significance. *Qualitative Phase:* It began with data familiarization, where audio recordings were transcribed verbatim and then translated from Swahili to English. A code book was developed to identify domains and generate initial codes. NVivo software was used to manage and analyze the data, facilitating cross-referencing of the 13 transcripts and organizing codes. Each code was thoroughly analyzed to capture the context and narrative of interviewees, providing a deep understanding of the facilitators and barriers related to adherence.

### Ethical approval and informed consent

Permissions were granted by the health facilities involved. Ethical clearance was secured from the MUHAS Institutional Review Board (Reference No. DA.282/298/01.C/2156). All participants provided written informed consent, which detailed the study’s objectives, as well as information on data privacy and confidentiality. Participants who did not receive services in line with the standard guidelines were counseled to address the discrepancies.

## Results

### Quantitative findings

#### Socio-demographic characteristics of participants

A total of 478 participants were recruited for this study, with 351 (73.43%) being female. The mean age of the participants was 42 years (SD: 12.06). Most participants, 158 (33.1%), were aged 45-54 years. Additionally, 278 (58.16%) of the participants resided in urban areas, and 154 (32.22%) were married. A significant portion, 319 (66.74%), had a primary education, and over the past 12 months, 393 (82.22%) reported having an income source “Table 1”

**Table 1:** Socio-Demographic Characteristics of Participants (n=478).

| Variable | Category | Frequency | Percentage (%) |
| --- | --- | --- | --- |
| Gender | Female | 351 | 73.43 |
|  | Male | 127 | 26.57 |
| **Age** | 15-24 years | 27 | 5.65 |
|  | 25-34 years | 87 | 18.20 |
|  | 35-44 years | 141 | 29.50 |
|  | 45-54 years | 158 | 33.05 |
|  | 55-64 years | 50 | 10.48 |
|  | 65+ years | 15 | 3.14 |
| **Residence** | Suburban | 200 | 41.84 |
|  | Urban | 278 | 58.16 |
| **Marital Status** | Widowed | 61 | 12.76 |
|  | Married | 154 | 32.22 |
|  | Living Together | 55 | 11.51 |
|  | Divorced/Separated | 105 | 21.97 |
|  | Never Married | 103 | 21.55 |
| **Income Source** | No | 85 | 17.78 |
|  | Yes | 393 | 82.22 |
| **Education Level** | Higher Education | 19 | 3.97 |
|  | Secondary Education | 89 | 18.62 |
|  | Primary Education | 319 | 66.74 |
|  | No Education | 51 | 10.67 |
| **Level of Facility** | Referral Hospital | 58 | 12.13 |
|  | Hospital | 40 | 8.37 |
|  | Health Center | 124 | 25.94 |
|  | Dispensary | 256 | 53.56 |
| **Profession (Cadre)** | Medical Doctor | 185 | 38.7 |
|  | Clinical Officer | 196 | 41 |
|  | Nurse | 97 | 20.29 |
| **Ever done CD4 test** | No | 35 | 7.43 |
|  | Yes | 443 | 92.68 |

#### Characteristics of selected ctc clinics

The study included 25 Care and Treatment Clinics (CTCs) in Dar es Salaam. Among these, dispensaries were 15(60%), with a mean volume of 2322 clients (SD: 2097.3). Additionally, 19(76%) of the clinics had at least minimum required number of staffs allocated. However, 16(64%) did not have staff who had received training. Furthermore, 15(60%) of the facilities did not receive any support supervision, and among those received supervision 15(60%) reported that did not receive feedback. The majority of the clinics, 24(96%) of the clinics had an assigned weighing scale, and 23(92%) had an assigned stadiometer. Moreover, 17(68%) did not have a MUAC tape “Table 2”.

**Table 2:** Characteristics of Selected CTC Clinics in Dar es Salaam (n=25)

| Variable | Category | Frequency | Percentage (%) |
| --- | --- | --- | --- |
| **Type of Health Facility** | Referral Hospital | 3 | 12 |
|  | Hospital | 2 | 8 |
|  | Health Center | 5 | 20 |
|  | Dispensary | 15 | 60 |
| **Number of Staff Allocated** | Inadequate | 6 | 24 |
|  | Adequate | 19 | 76 |
| **With Trained Staff** | No | 16 | 64 |
|  | Yes | 9 | 36 |
| **Received Support Supervision** | No | 15 | 60 |
|  | Yes | 10 | 40 |
| **Received Feedback After Supervision** | Don't Know | 2 | 8 |
|  | No | 15 | 60 |
|  | Yes | 8 | 32 |
| **Weighing Scale Available** | Yes, Assigned | 24 | 96 |
|  | Yes, Shared with other clinics | 1 | 4 |
|  | No | 0 | 0 |
| **Stadiometer Available** | No | 1 | 4 |
|  | Yes, Assigned | 23 | 92 |
|  | Yes, Shared with other clinics | 1 | 4 |
| **MUAC Available** | No | 17 | 68 |
|  | Yes, Assigned | 7 | 28 |
|  | Yes, Shared with other clinics | 1 | 4 |

#### Adherence of guideline in selected CTCs

Out of the 478 participants, 93 (19.46%) PLHIV received treatment which has fully adhere to the national nutrition guidelines. While follow-up care and anthropometric assessments were universally implemented (100%), the provision of micronutrient supplements was significantly lacking, with only 1% of participants receiving them “Figure 1”.

**Figure 1.**
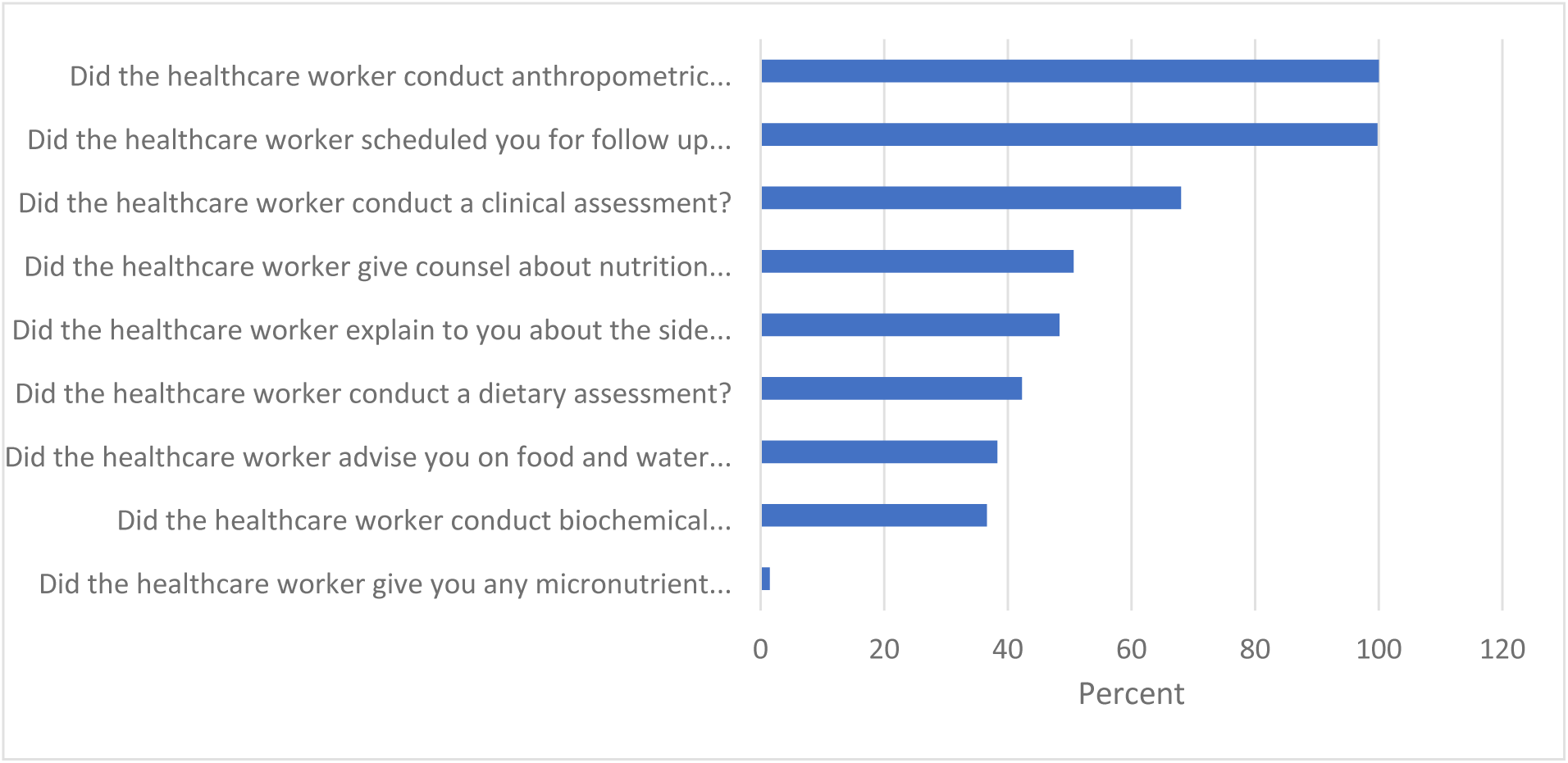
Adherence to Specific Aspects of Nutritional Care Guidelines.

Adherence rates to guidelines varied across different types of healthcare facilities. Referral hospitals had the highest adherence rate at 40%, indicating a strong compliance with the guidelines. Hospitals and health centers both showed moderate adherence rates, each at 25%. Dispensaries had the lowest adherence rate at 10%, suggesting potential challenges in maintaining guideline compliance in these facilities “figure 2”.

**Figure 2:**
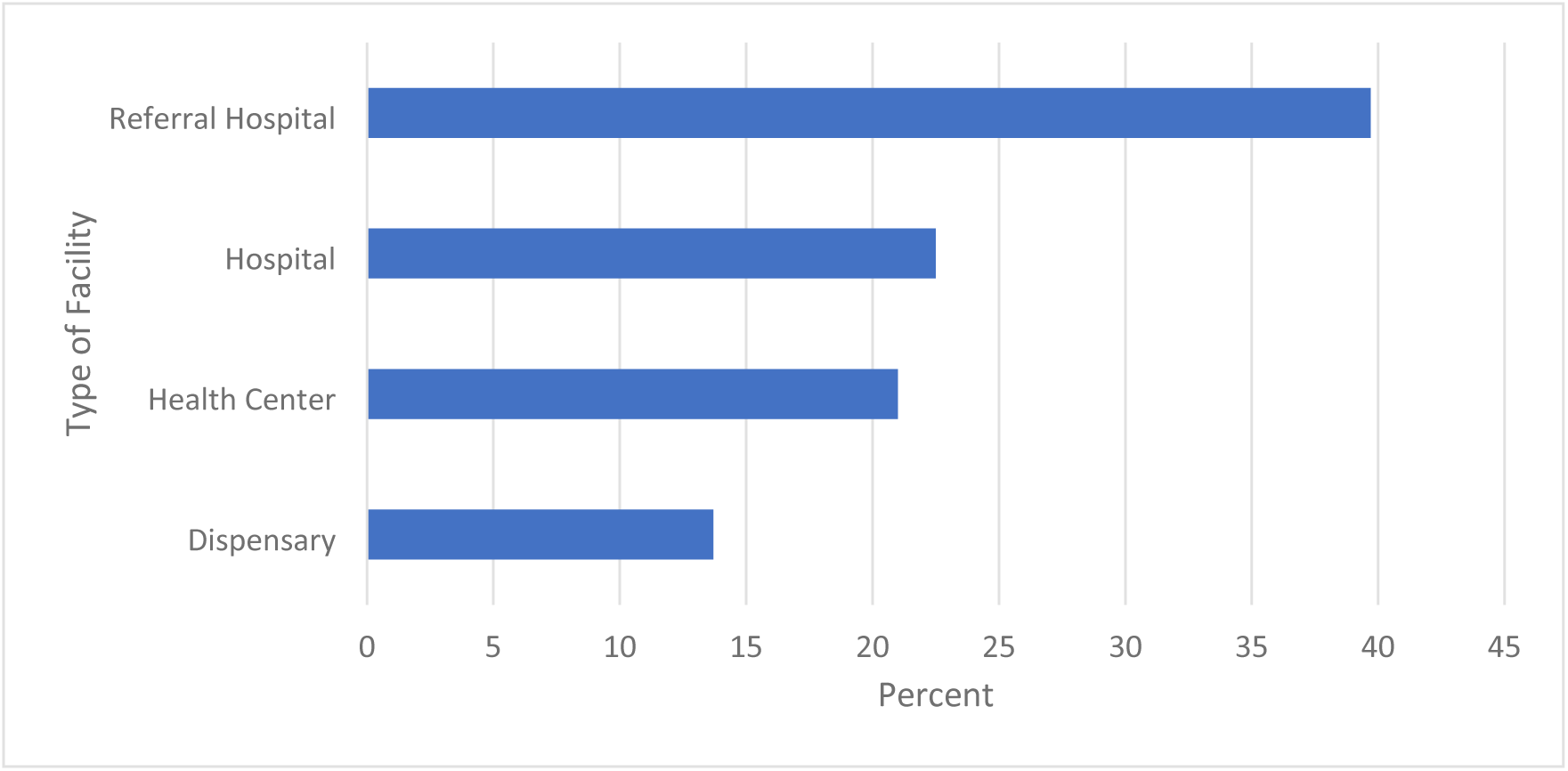
Adherence to the level of the facilities.

#### Determinants of adherence to guideline

In bivariate analysis, residence emerged as a significant determinant of adherence to the National Guidelines for Nutritional Care and Support. Higher adherence was observed in suburban areas 24%, (n=48) compared to urban areas 16.2%, (n=45) (χ2 = 4.5309, p = 0.033).

During multivariate analysis, several factors were identified as influencing adherence to nutritional guidelines among participants. Clients with higher education were significantly more likely to receive care that adhered to the National Guidelines for Nutrition Care and Support, with a 5.08 times higher adherence rate compared to those with no education (AOR=5.08; 95% CI: 1.30-19.79; p=0.019). Additionally, clients attending referral hospitals had a notably higher likelihood of receiving care that adhered to the National Guidelines, with an 8.23 times higher adherence rate compared to those attending dispensaries (AOR=8.23; 95% CI: 1.19-56.59; p=0.032).

Conversely, participants who were attended by nurses were less likely to receive care that adhered to the National Guideline for Nutrition Care and Support compared to those attended by doctors, with an AOR of 0.34 (95% CI: 0.70-4.42; p=0.038). Adequate staffing was associated with a lower likelihood of receiving care that adhered to the National Guideline for Nutrition Care and Support compared to having inadequate staffing, with an AOR of 0.31 (95% CI: 0.13-0.76; p=0.010). Urban residents were also less likely to receive care that adhered to the Nutrition guideline compared to those in sub-urban areas (AOR=0.47; 95% CI: 0.27-0.82; p=0.009) “Table 3”.

**Table 3.** Multivariate Logistic Regression Analysis of Factors Associated with Adherence.

| Variable Categories | Total (n) | COR (95% CI) | P-Value | AOR (95% CI) | P-Value |
| --- | --- | --- | --- | --- | --- |
| **Age Category** |  |  |  |  |  |
| **15-24** | 27 | 1 |  | Ref |  |
| **25-34** | 87 | 0.78(0.27-2.26) | 0.66 | 1.30(0.38-0.43) | 0.671 |
| **35-44** | 141 | 0.68(0.24-1.87) | 0.45 | 0.99(0.29-3.39) | 0.992 |
| **45-54** | 158 | 0.95(0.35-2.56) | 0.935 | 1.45(0.43-4.93) | 0.545 |
| **55-64** | 50 | 0.87(0.27-2.74) | 0.819 | 1.14 (0.29-4.53) | 0.845 |
| **65+** | 15 | 1.2(0.29-5.48) | 0.746 | 0.74(0.13-4.19) | 0.740 |
| **Gender** |  |  |  |  |  |
| **Female** | 351 | 1 |  | Ref |  |
| **Male** | 127 | 0.95(0.56-1.59) | 0.853 | 0.82(0.45-1.50) | 0.539 |
| **Education Level** |  |  |  |  |  |
| **No education** | 51 | 1 |  | Ref |  |
| **Primary** | 319 | 1.10(0.51-2.3) | 0.803 | 1.42(0.59-3.40) | 0.423 |
| **Secondary** | 89 | 1.02(0.41-2.51) | 0.961 | 1.40(0.50-3.89) | 0.511 |
| **Higher** | 19 | 2.72(0.83-8.83) | 0.096 | 5.08(1.30-19.79) | 0.019 |
| **Marital Status** |  |  |  |  |  |
| **Divorced** | 105 | 1 |  | Ref |  |
| **Living together** | 55 | 1.5(0.63-3.53) | 0.354 | 2.03(0.79-5.21) | 0.141 |
| **Married** | 154 | 1.45(0.73-2.85) | 0.280 | 1.91(0.90-4.04) | 0.088 |
| **Never Married** | 103 | 1.725(0.84-3.53) | 0.136 | 2.03(0.88-4.64) | 0.093 |
| **Widowed** | 61 | 1.78(0.79-4.01) | 0.160 | 1.74(0.70-4.32) | 0.227 |
| **Residence area** |  |  |  |  |  |
| **Sub-urban** | 200 | 1 |  | Ref |  |
| **Urban** | 278 | 0.61(0.38-0.96) | 0.034 | 0.47(0.27-0.82) | 0.009 |
| **Source income** |  |  |  |  |  |
| **No** | 85 | 1 |  | Ref |  |
| **Yes** | 393 | 0.74(0.422-1.30) | 0.297 | 0.84(0.42-1.67) | 0.628 |
| **Type of Facility** |  |  |  |  |  |
| **Dispensary** | 256 | 1 |  | Ref |  |
| **Health Center** | 124 | 1.67(0.95-2.93) | 0.071 | 0.88(0.43-1.79) | 0.737 |
| **Hospital** | 40 | 1.83(0.80-4.17) | 0.149 | 1.13 (0.37-3.42) | 0.828 |
| **Referral Hospital** | 58 | 4.14(2.19-7.83) | 0.000 | 8.23(1.19- 56.59) | 0.032 |
| **Provider Type** |  |  |  |  |  |
| **Clinical Officer** | 196 | 1 |  | Ref |  |
| **Nurse** | 97 | 0.33(1.13-0.84) | 0.020 | 0.34(0.70-4.42) | 0.038 |
| **Doctor** | 185 | 2.24(1.37-3.66) | 0.001 | 3.77(1.72-8.27) | 0.001 |
| **Staff Level** |  |  |  |  |  |
| **Inadequate** | 6 | 1 |  | Ref |  |
| **Adequate** | 19 | 0.82(0.47-1.44) | 0.500 | 0.31(0.13-0.76) | 0.010 |
| **Trained Staff** |  |  |  |  |  |
| **No** | 16 | 1 |  | Ref |  |
| **Yes** | 9 | 2.30(1.33-3.96) | 0.003 | 0.23(0.04-1.34) | 0.103 |
| **Supervision Status** |  |  |  |  |  |
| **No** | 15 | 1 |  | Ref |  |
| **Yes** | 10 | 2.09(1.32-3.30) | 0.002 | 0.69 (0.34-1.41) | 0.319 |
| **Tool (MUAC)** |  |  |  |  |  |
| **No** | 17 | 1 |  |  |  |
| **Yes, assigned** | 7 | 2.59(1.61-4.18) | 0.000 | 1.70(0.955-3.04) | 0.071 |
| **Yes, shared** | 1 | 1.07(0.30-3.84) | 0.908 |  |  |

### Quantitative Findings

Facilitators to adhering to guideline were training and competencies, organizational support, and patient education. Barriers included resource limitations, staff turnover, and documentation challenges “Table 4”.

**Table 4:**
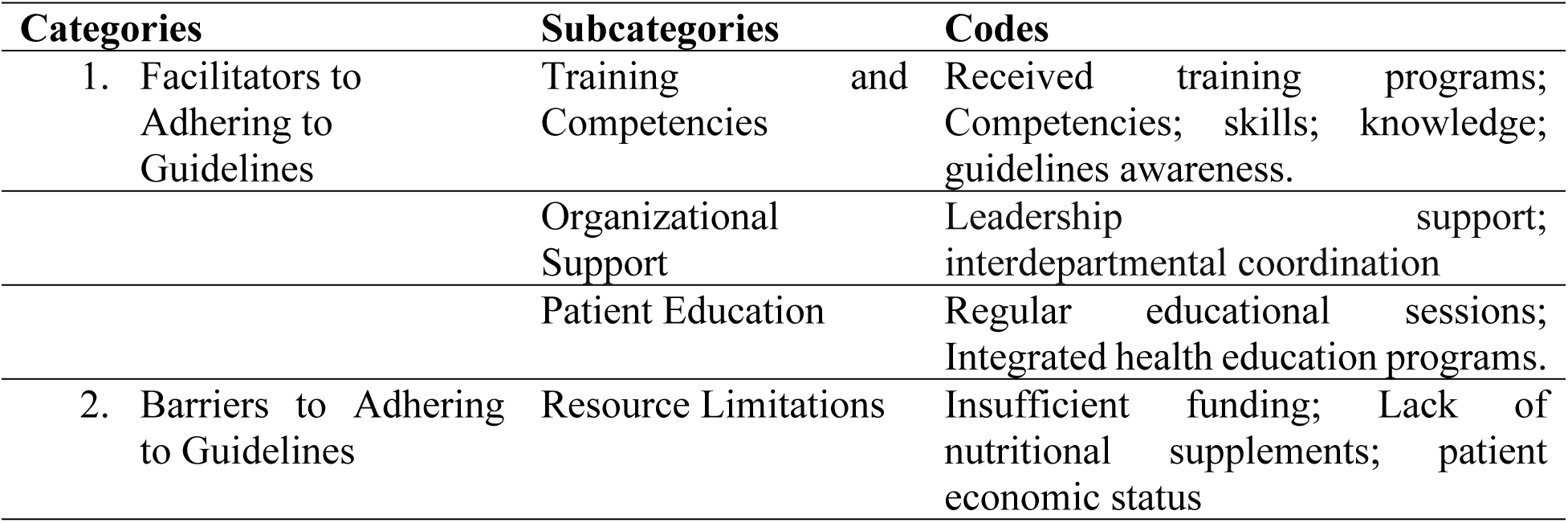
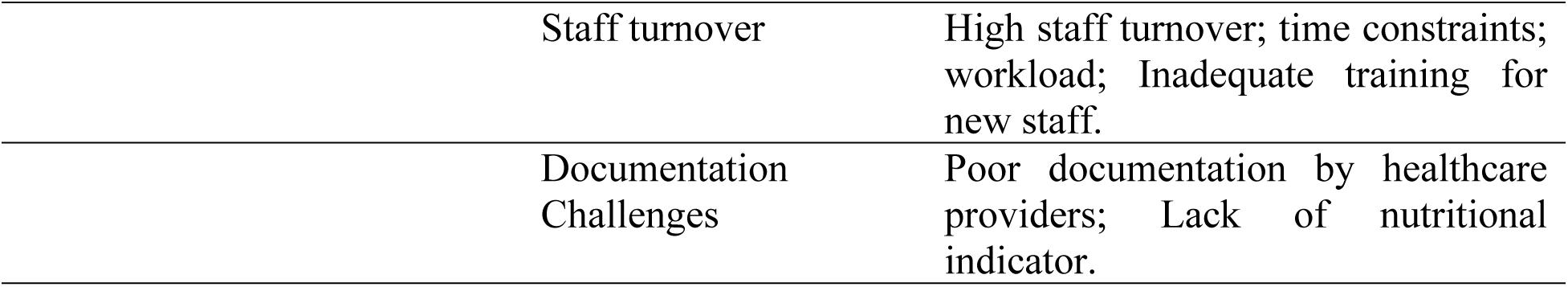
Facilitators and barriers in adhering to the guideline.

#### Training and Competencies

In this study, healthcare providers understand the importance of adhering to national nutritional guidelines for PLHIV. Their knowledge and abilities have improved greatly as a result of the training programs offered by different organizations.

*“I have attended around ten to fifteen nutrition training sessions related to the national nutrition guidelines Training has significantly improved my skills, they have significantly improved my competency, especially those from MUHAS we understand many things now…* (IDI participant 13) Organizational support and Guideline.

Participants pointed that effective leadership has shown a crucial role in ensuring adherence to nutritional guidelines, particularly in the context of managing HIV. It involves not just supervision but also fostering a supportive and motivating environment for staff to consistently meet established standards.

*“The specific ways our leaders use involve internal supervision, which is conducted to monitor the clients served throughout the week. This is because there is something called ’backup.’ When this backup is sent, it goes primarily to the Medical Officer In charge, who reviews it since they are the leaders, we have here…”* (IDI participant 3)

This proactive leadership ensures that staff are well-informed about guidelines and receive ongoing support and oversight, promoting a culture of continuous improvement.

*“Our leaders remind us every day to follow the guidelines. They provide us with the guidelines, which we read regularly. We also have weekly meetings where they guide us on following the guidelines……They help by providing tools designed to guide us, which they regularly monitor these tools to ensure we are following the guidelines. They check and review the tools, which helps remind us of anything we might have forgotten.”* (IDI participant 7)

#### Patient Education

Participant explained that Regular educational sessions are essential for enhancing patients’ understanding of nutrition, especially for those with HIV. These sessions offer ongoing, organized opportunities to learn about proper nutrition and its significance in managing overall health.

*“We usually provide education every morning before starting services at 7:30 am. The education we provide covers many topics, including nutrition, because the medication they take requires food. Without sufficient education, people cannot understand this, so we emphasize the importance of food, the importance of leafy vegetables, lifestyle nutrition, and which foods are not good for health.”* (IDI participant 9)

#### Resource Limitation

Nutritional supplements have been inconsistent, often reliant on fluctuating external funds, this leads to period of unavailability of essential nutritional supplements, which it negatively impacts the health and nutrition of PLHIV.

*“Unfortunately, we’re facing challenges. We lack therapeutic and supplemental foods, which hinders our progress. A client comes in with needs, really needs nutritional supplements. So, just talking to them and telling them to do one, two, three things set us back. It’s frustrating; there’s no perfect way to express this”.* (IDI participant 8)

#### Staff Turnover

Participant pointed out staff turnover in healthcare, especially in complex care settings like HIV treatment, compromises patient care by disrupting continuity and overburdening remaining staff.

“*When you talk about the reduction in healthcare providers going on leave, it significantly affects service delivery because, for example, here at Goba, we have very few staff members. So, when one staff member leaves, the workload becomes much heavier, especially in serving our clients…. Therefore, staff turnover impacts our adherence to guidelines because you won’t be able to do everything that is required for the patient.”* (IDI participant 3)

#### Documentation Challenges

Poor documentation and the absence of clear nutritional care standards are critical gaps compromising healthcare quality. Incomplete records and lack of specific indicators hinder effective patient care and outcomes.

*“Those of us working in the CTC have indicators, so our major task is to engage with those indicators…… We concentrate on working with these indicators. For example, for viral load suppression, we implement all mechanisms to ensure it increases… However, for nutrition, we don’t have such an indicator, which is a challenge* ”(IDI participant 4)

## Discussion

Only nineteen percent of PLHIV received care which fully adhered to the National Nutrition Guidelines, a rate significantly lower than regions like Ethiopia where adherence ranges from 36.3% to 41% [20]. The universal implementation of follow-up care and anthropometric assessments reflects a commitment to some aspects of nutritional care, but highlights insufficiencies in holistic patient management, echoing WHO recommendations for comprehensive care [21, 22] . Only one percent of participants received essential micronutrient supplements, indicative of systemic incapacities including supply chain failures. This deficiency has significant implications for immune function and health in HIV-infected individuals, underscoring a gap that is starkly highlighted by WHO’s emphasis on the importance of these nutrients [23]. Clinical nutritional assessments were performed for sixty percent of participants, suggesting moderate adherence. However, less than half received dietary assessments, side effects monitoring, hygiene assessments, and nutrition counseling, pointing to significant gaps in nutritional management. These findings are consistent with higher rates of dietary assessments and nutrition counseling reported in studies from Kenya and Uganda, suggesting a regional disparity in adherence practices [24, 25].

Educational level was a prominent determinant of adherence, with individuals having higher education showing significantly better compliance, underlining the importance of health literacy in guideline adherence. This finding is supported by a study in Ghana, showing that education correlates strongly with better nutritional outcomes [26]. The type of healthcare facility also critically influenced adherence, with referral hospitals achieving the highest rates (40%) due to better resources and specialized staffing, contrasting sharply with dispensaries that showed only 10% adherence, highlighting the resource disparity across facility levels. This pattern is echoed in Malawi, where private health facilities noted higher adherence rates [27]. Provider type was another key factor, with doctors achieving higher adherence in their care compared to nurses, suggesting that provider training and qualifications play significant roles in guideline adherence. Resource availability within healthcare facilities also had a substantial impact, with well-resourced facilities demonstrating higher adherence, similar to findings from Mozambique where better-equipped facilities saw more frequent patient visits [28].

Consistent training was reported to be facilitator that linked to improved healthcare delivery, similar to findings by Sunguya et al. (2013), which showed significant enhancements in health workers’ nutrition knowledge post-training [29]. Strong organizational leadership and robust feedback mechanisms, was identified as another facilitator, enhancing adherence by enabling prompt corrections to deviations from protocols, echoing a study in Mozambique that highlighted the impact of healthcare service availability on care-seeking behaviors. Interdepartmental coordination was essential for integrated patient care, especially for HIV patients requiring multifaceted approaches, aligning with the Mozambican study’s emphasis on facility type and resource levels influencing healthcare decisions [28]. Patient education was pivotal in improving health outcomes, supported by findings from Nigeria showing higher adherence rates among patients educated in private health facilities [30].

Conversely, significant barriers included resource limitations, particularly nutritional supplements, which directly affected care quality, a challenge also noted in Mozambique [28]. Staffing turnover and shortages posed substantial barriers, increasing workloads and potentially leading to caregiver burnout, necessitating strategic workforce planning to maintain care continuity, a strategy supported by findings from Uganda [31, 32]. Documentation challenges impaired nutritional care quality, where poor practices hindered effective intervention monitoring, a situation improved by standardized documentation practices as shown in studies by Kight et al. (2019) advocating for electronic health records in the country [33]. Economic constraints also critically impacted patients’ adherence to nutritional guidelines, with financial hardships forcing compromises in dietary quality, a dynamic observed in study by Correia (2017) on the interplay between economic conditions and healthcare outcomes [34]. Integrated support services providing both nutritional and economic assistance were deemed vital for enabling patients to adhere to dietary recommendations effectively as supported by study by (Håkonsen et al., 2019) [35, 36].

External validity of findings is limited by the unique healthcare infrastructure and socio economic context of Dar es Salaam; hence they may not accurately represent the conditions in less developed regions. Additionally, response bias is a concern, as data from patients and healthcare providers were partially self-reported, the inclusion of an observational component of the study helped to mitigate this bias.

## Conclusion

Our study found a significant gap in adherence to national nutritional guidelines among HIV patients, with only nineteen percent receiving care that fully adhered to the national nutritional guideline, much lower than similar studies elsewhere. This underscores a critical need for improvement. While follow-up care and anthropometric assessments were robustly implemented, a severe shortfall in essential micronutrient supplements suggests systemic failures, potentially involving supply chain and funding issues. Variations in adherence across different healthcare facilities indicate that resource distribution, staff training, and facility capabilities are crucial for guideline compliance. Socio-demographic factors such as education levels and geographical location also significantly influenced adherence outcomes. Enhancing training for healthcare providers through continuous, independent programs to maintain high standards of care, strengthening resource allocation to lower-tier health facilities, and expanding patient education to improve engagement and adherence. Additionally, addressing systemic barriers to ensure consistent supply of essential supplements and improving documentation practices are essential for effective patient management and improved health outcomes.

## Data Availability

Data generated and analysed from this study are available upon a reasonable request to the corresponding author.

## Acknowledgement

The authors wish to acknowledge dedicated academic staff at the School of Public Health, and the Department of Development Studies at MUHAS, the Regional AIDS Control Coordinator office and the CTC clinics in Dar es salaam whose organized efforts, professional guidance, and unwavering support created a research-friendly environment that was crucial to the conceptualization and implementation of this study.

## References

[1] World Health Organization. Global progress reports.

[2] Singhato A, Khongkhon S, Rueangsri N, et al. Effectiveness of Medical Nutrition Therapy to Improve Dietary Habits for Promoting Bone Health in People Living with Chronic HIV. Ann Nutr Metab 2021; 76: 313–321.

[3] United Nation programme on HIV/AIDS. Global HIV statistics People living with HIV People living with HIV accessing antiretroviral therapy New HIV infections AIDS-related deaths.

[4] Seid A, Seid O, Workineh Y, et al. Prevalence of undernutrition and associated factors among adults taking antiretroviral therapy in sub-Saharan Africa: A systematic review and meta-analysis. PLoS One; 18. Epub ahead of print 2023. DOI: 10.1371/journal.pone.0283502.

[5] John-Joy Owolade A, Abdullateef RO, Adesola RO, et al. Malnutrition: An underlying health condition faced in sub Saharan Africa: Challenges and recommendations. Annals of Medicine and Surgery; 82. Epub ahead of print 2022. DOI: 10.1016/j.amsu.2022.104769.

[6] Tanzania Food and Nutrition Centre (TFNC). National Guidelines For Nutrition Care And Support OF People With HIV. 2016.

[7] Ndakala FN, Oyugi JO, Oluka MO. HIV-Associated Polyneuropathy in Resource-Limited Settings: Genetic Predisposition and Vitamin Variations. World J AIDS 2017; 07: 106–121.

[8] Weiser SD, Young SL, Cohen CR, et al. Conceptual framework for understanding the bidirectional links between food insecurity and HIV/AIDS. American Journal of Clinical Nutrition 2011; 94: 1729S–1739S.

[9] Kabalimu TK, Sungwa E, Lwabukuna WC. Malnutrition and associated factors among adults starting on antiretroviral therapy at PASADA Hospital in Temeke District, Tanzania. Tanzan J Health Res; 20.

[10] Takarinda KC, Mutasa-Apollo T, Madzima B, et al. Malnutrition status and associated factors among HIV-positive patients enrolled in ART clinics in Zimbabwe. BMC Nutr 2017; 3: 1–11.

[11] Fathima AS, Madhu M, Udaya Kumar V, et al. Nutritional Aspects of People Living with HIV (PLHIV) Amidst COVID-19 Pandemic: an Insight. Curr Pharmacol Rep 2022; 8: 350– 364.

[12] Delisle H, Shrimpton R, Blaney S, et al. Capacity-building for a strong public health nutrition workforce in lowresource countries. Bull World Health Organ 2017; 95: 385– 388.

[13] NACP. Tanzania Hiv Impact Survey (This) 2016-2017. Tanzania HIV Impact Survey (THIS) 2016-2017 2018; 2016–2017.

[14] Kabalimu T, Sungwa E, Lwabukuna W. Malnutrition and associated factors among adults starting on antiretroviral therapy at pasada hospital in Temeke district, Tanzania. Tanzan J Health Res; 20.

[15] Heath K, Levi J, Hill A. The Joint United Nations Programme on HIV/AIDS 95-95-95 targets: worldwide clinical and cost benefits of generic manufacture. Aids 2021; 35: S197–S203.

[16] Aber H, Kisakye AN, Babirye JN. Adherence to complementary feeding guidelines among caregivers of children aged 6-23 months in Lamwo district, rural Uganda. Pan African Medical Journal 2018; 31: 1–11.

[17] Golley RK, Smithers LG, Mittinty MN, et al. An index measuring adherence to complementary feeding guidelines has convergent validity as a measure of infant diet quality. Journal of Nutrition 2012; 142: 901–908.

[18] TDHS. Demographic and Health Survey and Malaria Indicator Survey. Paper Knowledge Toward a Media History of Documents 2022; 1–23.

[19] USAID’s SPRING Project. Tool for Rapid Evaluation of Facility-Level Nutrition Assessment, Counseling, and Support: A User’s Guide.

[20] MoH. National Guidelines for HIV/AIDS and Nutrition in Ethiopia.

[21] WHO. Nutritional care and support for people living with HIV/AIDS A training course.

[22] Mirzazadeh A, Eshun-Wilson I, Thompson RR, et al. Interventions to reengage people living with HIV who are lost to follow-up from HIV treatment programs: A systematic review and meta-analysis. PLoS Med 2022; 19: e1003940.

[23] WHO Library Cataloguing-in-Publication Data WHO Technical Consultation on Nutrient Requirements for People Living with HIV/AIDS (2003 : Geneva, Switzerland) Nutrient requirements for people living with HIV/AIDS : report of a technical consultation.

[24] Mwangi A. The Effects of Nutritional Knowledge on the Dietary Practices of People Living with HIV in Kayole Division, Nairobi-Kenya. International Journal of Nutrition and Food Sciences, https://www.academia.edu/46977136/The_Effects_of_Nutritional_Knowledge_on_the_Dietary_Practices_of_People_Living_with_HIV_in_Kayole_Division_Nairobi_Kenya (2014, accessed 17 June 2024).

[25] González-Alcaide G, Menchi-Elanzi M, Nacarapa E, et al. HIV/AIDS research in Africa and the Middle East: Participation and equity in North-South collaborations and relationships. Global Health 2020; 16: 1–18.

[26] Nanewortor BM, Saah FI, Appiah PK, et al. Nutritional status and associated factors among people living with HIV/AIDS in Ghana: cross-sectional study of highly active antiretroviral therapy clients. BMC Nutr 2021; 7: 1–8.

[27] Chirambo L, Valeta M, Banda Kamanga TM, et al. Factors influencing adherence to antiretroviral treatment among adults accessing care from private health facilities in Malawi. BMC Public Health 2019; 19: 1–11.

[28] Anselmi L, Lagarde M, Hanson K. Health service availability and health seeking behaviour in resource poor settings: evidence from Mozambique. Health Econ Rev 2015; 5: 1–13.

[29] Sunguya BF, Poudel KC, Mlunde LB, et al. Nutrition training improves health workers’ nutrition knowledge and competence to manage child undernutrition: A systematic review. Front Public Health 2013; 1: 63771.

[30] Monjok E, Smesny A, Okokon IB, et al. Adherence to antiretroviral therapy in Nigeria: an overview of research studies and implications for policy and practice. HIV/AIDS - Research and Palliative Care 2010; 2: 69–76.

[31] Lugada E, Ochola I, Kirunda A, et al. Health supply chain system in Uganda: assessment of status and of performance of health facilities. J Pharm Policy Pract; 15. Epub ahead of print 1 December 2022. DOI: 10.1186/S40545-022-00452-W.

[32] MoH. Uganda Services Availability and Readiness Assessment 2013 Summary Report: Key findings in figures Uganda 2013 Service Availability and Readiness (SARA) Index GENERAL SERVICE READINESS.

[33] Adeniyi AO, Arowoogun JO, Chidi R, et al. The impact of electronic health records on patient care and outcomes: A comprehensive review. https://wjarr.com/sites/default/files/WJARR-2024-0592.pdf 2024; 21: 1446–1455.

[34] Raji RO. Nutrition Intake, Health Status, Education and Economic Growth: A Causality Investigation. Econometric Research in Finance 2020; 5: 79–102.

[35] SNAP Is Linked With Improved Health Outcomes and Lower Health Care Costs | Center on Budget and Policy Priorities, https://www.cbpp.org/research/food-assistance/snap-is-linked-with-improved-health-outcomes-and-lower-health-care-costs (accessed 17 June 2024).

[36] Behrman R. THE IMPACT OF HEALTH AND NUTRITION ON EDUCATION.

